# A global analysis of domestic military policies governing responses to public health emergencies

**DOI:** 10.1101/2024.10.12.24315372

**Authors:** Kuang Yu Hu, Ciara M. Weets, Rory Wilson, Gunnar V. Ljungqvist, Rebecca Katz

## Abstract

Throughout the COVID-19 pandemic, militaries around the world mobilized at an unprecedented scale to support domestic response efforts. This was consistent with the growing trend of asset mobilization for military operations other than war during public health emergencies. However, the global scale and vast breadth of civil-military cooperation during the pandemic invites new considerations regarding the authority and scope of domestic operations of militaries during public health emergencies. We have systematically analyzed domestic military deployment policies in each UN member state, focusing on the authority, execution and scope of military involvement pertaining to domestic public health emergencies.

We analyzed legally enforceable policies, including Constitutions, defense ministry authorizations, and legal frameworks. We categorized how each country codified the deployment of military assets, who holds authority for deployment and the procedural mechanisms for deployments. Our findings revealed that of countries with active military forces, nearly all (170/171) have codified rules on domestic military deployment and 90.59% (154/170) allow military mobilization through executive orders. Furthermore, 58.48% (100/171) of countries with an active military have codified separation of powers to ensure that civilian decision makers are exclusively empowered to mobilize military forces. Finally, we found that 74.85% (128/171) of countries included language that authorized military involvement in domestic military operations other than war.

Our findings provide critical data for analyzing the relationship between military operations and public health outcomes, including how specific domestic military deployment policies impact the speed and effectiveness of military involvement in public health emergencies.

## Introduction

During the COVID-19 pandemic, militaries around the world were mobilized at an unprecedented scale to support domestic outbreak response efforts. This was consistent with the growing trend of asset mobilization for military operations other than war (MOOTW), during national emergencies, especially humanitarian crises and outbreak responses [1]. However, the global scale and vast breadth of military involvement during the COVID-19 pandemic invites new considerations regarding the authority and scope of domestic operations of militaries during public health emergencies.

Civil-military cooperation describes instances where militaries cooperate with civilian authorities in MOOTW. Civil-military cooperation in public health responses involves using military resources and capabilities to provide clinical, logistical and personnel support to assist public health interventions [2]. Their ability to quickly organize large numbers of personnel, facilitate logistics and transport, and provide self-sustaining medical care is a unique capability that few civilian agencies can replicate [2]. Historically, militaries have significantly contributed to public health responses ranging from biomedical research, healthcare provision in conflict settings and implementing interventions during epidemics. However, over the past two decades militaries have increasingly been deployed domestically and internationally to provide humanitarian relief and assist in outbreak responses [3]. The growing role of military deployment in outbreak responses became particularly apparent during the 2014-2016 West Africa Ebola outbreak and the 2015-2016 Latin American Zika epidemic [4].

Civil-military cooperation became critical from the start of the COVID-19 pandemic, with an estimated 95% of militaries engaged in assisting their domestic public health response [5]. The nature of civil-military cooperation was varied due to differences in political systems, state leadership and political legacies. Gibson-Fall [6] proposed three distinct trends in civil-military cooperation during the first 6 months of the COVID-19 pandemic. These were minimal technical military support, blended civil-military cooperation responses and military-led responses.

The diversity of public health response roles that militaries performed worldwide during the COVID-19 pandemic is striking [7]. The pandemic massively accelerated and expanded the trend of deploying militaries in public health responses. This has led to important questions on current policies governing domestic deployment of militaries in public health emergencies. Therefore, we aimed to identify the domestic military deployment policies in each UN member state, and assess how each nation codifies the roles and responsibilities of militaries in domestic emergencies. We also sought to capture data that might impact outcomes, such as decision making authority for using a military domestically. These data will enable future research into how domestic civil-military cooperation policies impact the outcomes of outbreaks and natural disasters across diverse contexts.

## Methods

### Country Selection and Project Scoping

We analyzed legally enforceable policies, including Constitutions, defense ministry authorizations, and legal frameworks from 193 United Nations (UN) Member States. The Democratic People’s Republic of Korea was not included in this analysis, due to lack of access to relevant documents. All policies were identified, analyzed, and integrated into the customized data taxonomy between October 2023 and August 2024.

This military dataset was collected as part of the larger Analysis and Mapping of Policies for Emerging Infectious Diseases (AMP EID) project, which creates and analyzes repositories of policy documents. All policies are collected to inform decision making during outbreaks, understand the global regulatory environments with implications for infectious disease events, and to be incorporated into epidemiological modeling. Across the project, a single standardized operating procedure (SOP) was utilized to direct data collection. Projects began with a literature review to identify relevant boolean search terms to surface relevant policies for the topic. These search terms and the data collection methodology were then tested in a proof-of-concept study of ten countries. At the conclusion of the proof-of-concept study, the research team resolved methodological issues, finalized the customized search protocol and created a standardized coding procedure. The data taxonomy was then developed for data collection and coding.

### Policy Identification Protocols

Military deployment processes and authorities are often found in the constitution or other foundational legal documents of a nation. As such, we began data collection by surfacing and reviewing the contents of all digitized constitutions of the 193 UN Member States selected for study. As five countries do not have a formal constitution, they were excluded from this step.

We then completed a search of online legal repositories for each nation, reviewing any legally-enforceable policy with a title containing the terms ‘armed forces’, ‘defense’, ‘emergency’, ‘security’, ‘army’ or ‘military’. If the policy document cross-referenced another relevant document, that document was searched for by name and reviewed. To ensure comprehensive data collection, we finally conducted a series of standardized searches using boolean search terms (Table 1). In the case that the target country conducted government affairs in a language other than English, Google Translate was used to translate these search terms into the language primarily used to conduct and communicate governmental actions.

**Table 1.** Search term series employed in policy identification protocol.

| Sequence | Query Term |
| --- | --- |
| 1 | "[Country name] AND constitution" |
| 2 | "[Country name] AND "(defense OR "armed forces") act")" |
| 3 | "[Country name] AND "emergency act" |

### Policy Analysis

Potentially relevant policies were preliminarily screened to ensure that the identified version was current and legally-enforceable. As a result, strategies, reports, draft laws, and repealed laws were excluded, as these documents are not enforceable within a country’s legal system. All screened policies were then downloaded as PDFs, and captured in Airtable, a cloud-based platform that hosts relational databases.

For policies in languages not spoken by any research team members, machine translation was used to complete translation of policies into English. When possible, fluent speakers of these languages were contacted by the research team to verify the translation of relevant policy provisions.

For all policies that passed preliminary screening, the research team completed comprehensive reviews of their contents, assessing them against the standardized inclusion criteria (Supplementary Information 1). Policies that met inclusion criteria were then entered into the customized data taxonomy and countries were assigned to a qualitative category (‘status’) for each of the research categories (‘subtopics’) associated with the research questions (Figure 1).

**Figure 1.**
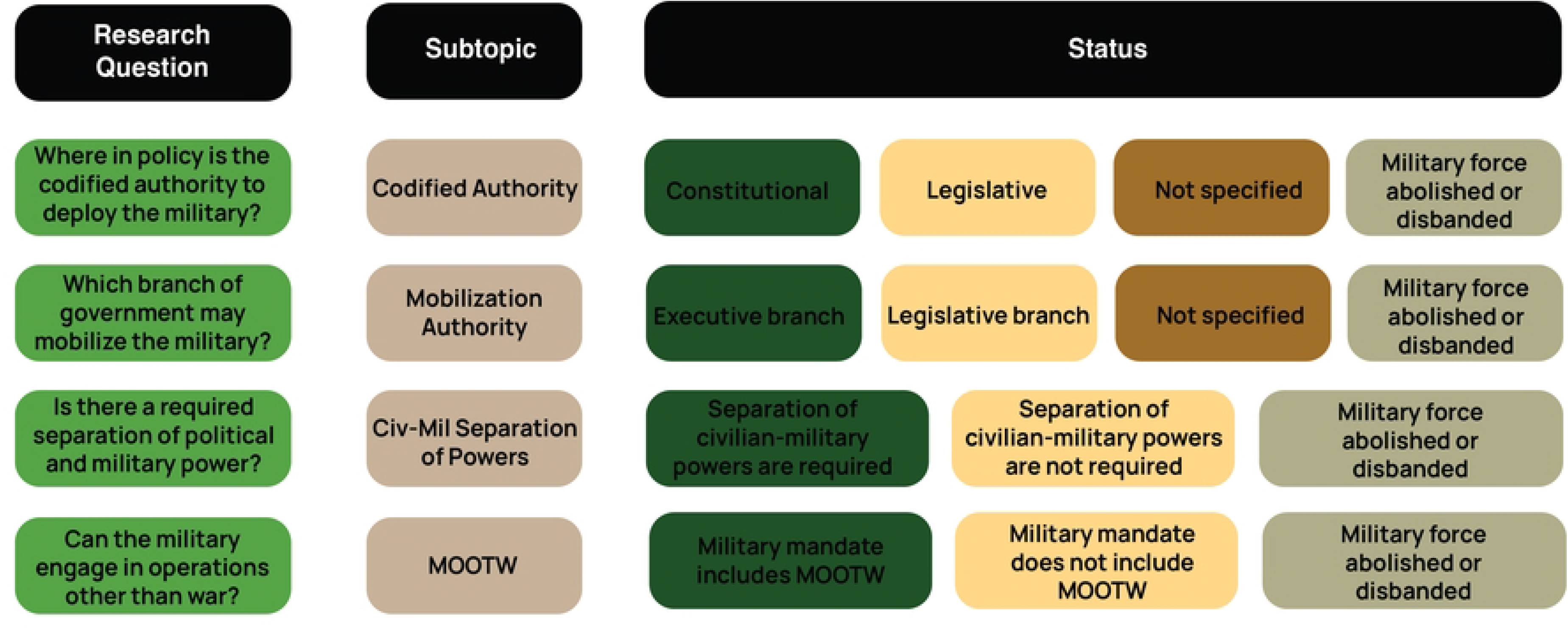
Customized data taxonomy for military engagement policy data collection. Figure should be read from left to right. Each research question corresponds to the subtopic and status to its right.

The literature review, methodology, and inclusion criteria were approved by the entire research team and the Principal Investigator. To ensure uniform decision making, the research was initially conducted by a primary researcher. To validate the policy coding of the primary researcher, a second member of the research team then completed a review of policies and associated codes. Any coding discrepancies between the primary and secondary reviewer were deconflicted by the research team and the Principal Investigator. The full datasets, reproducible code, and figures are available at [8] [https://github.com/cghss/military_engagement].

## Findings

We found that 88.60% (171/193) of UN Member States have an active national military force, while the remaining 11.40% (22/193) of countries have had their militaries disbanded or abolished. These 22 nations without a military force were removed from subsequent calculations, as this research effort focused solely on countries that had active militaries. Of those nations that have an active military, nearly all (170/171) have codified rules on which decision makers may deploy armed forces and how such deployments of military assets are ordered. Jamaica does not define the triggering mechanism or process for military asset deployment and was therefore removed from subsequent calculations in these categories [9]. All countries (171/171) were found to define the scope of military operations, though qualitative analysis found that there is significant variation in how broad the mandate for military intervention is from country to country.

### Military Mobilization Processes for Domestic Deployments

Nearly all countries dictate the process by which national military forces are mobilized and deployed within either their constitution or legislation. Of the nations with a military, 77.65% (132/170) included the military mobilization processes in the constitution. Inclusion of these authorities was found to be commonplace, regardless of region or form of government. By contrast, 22.35% (38/170) of countries included this mobilization process in legislation (Figure 2). Five countries lack constitutions, but codify their mobilization processes and authorities in seminal legislation. This is the case in Saudi Arabia and New Zealand, neither of which have a formal constitution, but instead have a collection of formative legislation in which the legal processes for military deployments are documented [10,11].

**Figure 2.**
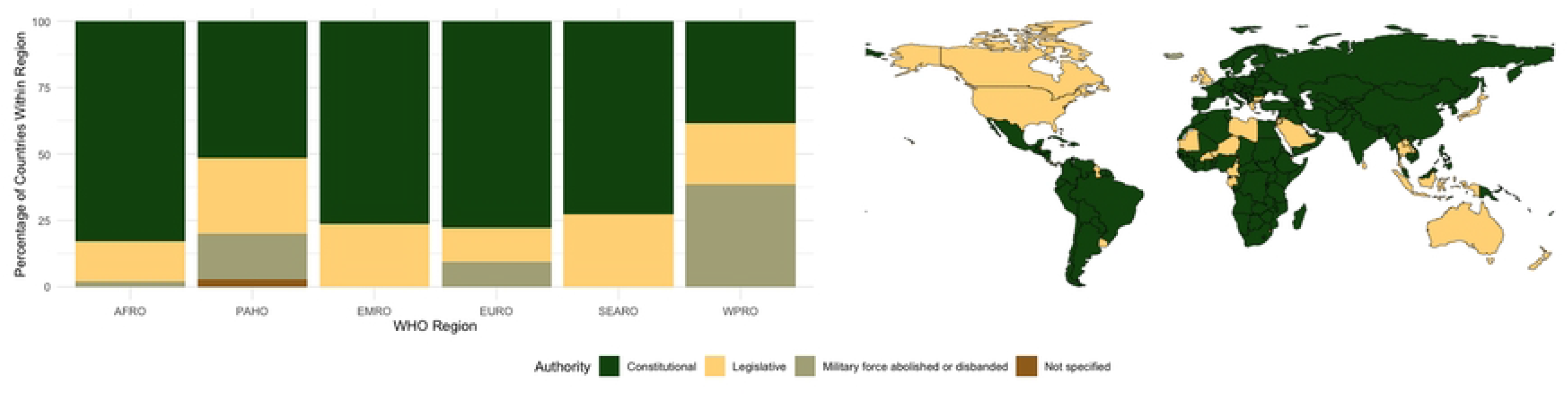
Regional and geographical distribution of policy location of codified military deployment process.

**Figure 3.**
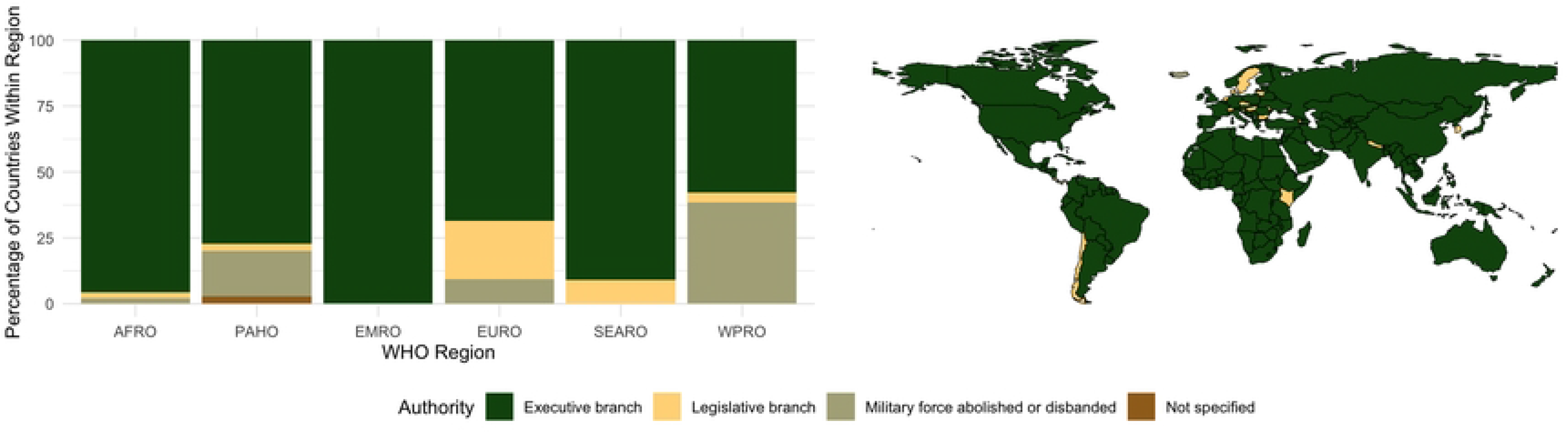
Regional and geographical distribution of governing bodies authorized to deploy military assets domestically.

**Figure 4.**
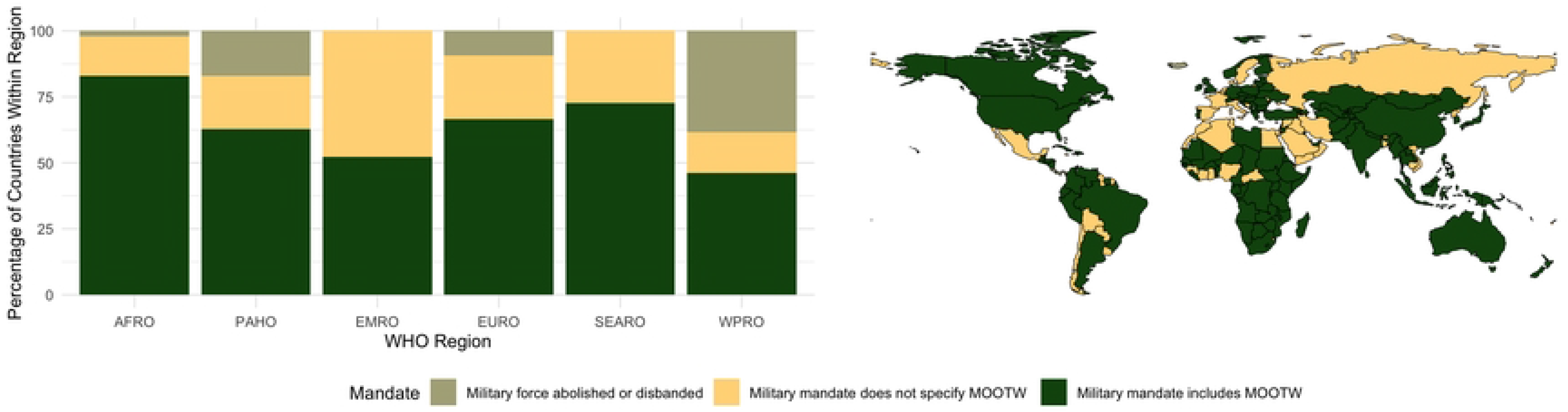
Regional and geographical distribution of permitted scope of domestic military engagement.

Policies documenting military mobilization processes universally include triggering mechanisms that initialize the cascade of events required for troop mobilization and deployment. We found that these triggering mechanisms always fell under either executive orders or legislative actions. The majority of countries with active militaries, 90.59% (154/170), mobilize military assets through an order from an executive decision maker. However, due to the diversity of global political systems, these executive orders may take different forms. For example, in Nigeria, military assets may be mobilized by order of the sitting president, whereas in Bhutan, military forces are deployed at the order of the King [12,13]. In 16 countries (9.36%), mobilization of the military cannot be authorized without approval of a legislative body, regardless of the request of the executive (Figure 2). For example, in the Republic of Korea, a presidential order to deploy the military requires approval from the national assembly. In Denmark, except in response to an invasion of the country, the King can only mobilize military forces with approval of the legislative body [14,15].

### Civil-Military Separation of Powers

The legally-enforceable policies and constitutions that dictate the process and mechanisms by which military assets may be deployed often also prohibit active military personnel from holding political roles in the government. These stipulations create a separation of powers between political and military leadership, and therefore, ensure that military mobilization orders come from civilian authorities. We found that 58.48% (100/171) of countries with an active military have codified such separation of powers to ensure that civilian decision makers are exclusively empowered to mobilize military forces. Notably, countries with histories of military coups in the modern era that have since adopted democratic practices, such as Argentina, Chad, and Chile have policies prohibiting active military personnel from holding executive office [16–18]. The remaining 41.52% (71/171) of nations with militaries allow active military personnel to hold executive offices. Often these countries have a constitutional monarchy system, such as the United Kingdom or Jordan, where the regent acts as the head of state, commands the military, and has political influence [19,20]. However, this is also seen in countries that exclusively concentrate political and military power under a single authority body or figure, such as the Islamic Republic of Iran and the Lao People’s Democratic Republic [21,22].

### Scope of Permissible Domestic Military Deployments

The scope of military engagement after deployment is often included in policies, though we found that it tends to be less well defined than the triggering mechanism and asset mobilization process. In order to capture the diversity of terms and authorities given to military assets to engage in domestic, non-combat missions, we intentionally employed the broad term ‘Military Operations Other Than War (MOOTW)’. 74.85% (128/171) included language that authorized military involvement in domestic MOOTW. Many times, these were explicit authorities, as in India, where military troops were authorized under the Disaster Management Act of 2005 to aid in domestic natural disaster response, states of emergency, and public health assistance [23]. In other cases, such as that of Peru, the constitution states that the military may be used to assist in cases of national emergency, which may be triggered by a disruption to the peace or public order due to catastrophe or grave circumstance [24]. Although Peru provides less prescribed circumstances for involvement of the military in domestic MOOTW relative to India, both countries could be construed to legally employ military assets in non-combat missions triggered by emergencies. The remaining 44 countries (25.73%) with active militaries do not include language in legally-enforceable policy to allow for domestic MOOTW. While some may include broad language, such as the Constitutions of Bolivia or Algeria, which states that troops may be involved in the defense of the state and its interests, there was no terminology included in the provision to definitively suggest that non-combat operations would be included in this mandate [25,26]. While countries that do not explicitly authorize MOOTW tend to be evenly distributed around the world, we found a noticeable concentration of countries in the World Health Organization’s Eastern Mediterranean Region (EMRO). Within EMRO, nearly half of the nations (47.62%; 10/21) do not have codified authorities for MOOTW, rendering it the region with the smallest proportion of countries that have policy permitting MOOTW.

## Discussion

In mapping the international policy landscape, we have established the authority, the execution, and the scope of domestic MOOTW. The authority of military deployment has been described as either constitutional or legislative, and whether it is under civilian control or not. The execution of domestic MOOTW has been codified as either legislative or executive, describing the two main processes by which the authority may deploy military forces for MOOTW. The scope of civil-military operations has also been described, and whether MOOTW were explicitly mandated or not in different countries. These data provide important context on the extent of military powers during MOOTW on civil society, and what guardrails are in place to prevent excessive force of militaries on its civilian population. This dataset thus presents several dimensions of civil-military operations policy as they may be potentially relevant for pandemic preparedness and response.

In the shadow of the COVID-19 pandemic, many questions remain unanswered regarding how countries deployed their militaries as part of the pandemic response. This paper thus offers critical information when analyzing the relationship between civil-military operations and public health outcomes. For instance, it is possible that specific policy patterns impact the speed or effectiveness with which countries were able to deploy their militaries. These include whether legislative approval delayed responses, whether allowing military officials to hold political office led to over-militarization of public health responses and whether a lack of clear MOOTW authorization delays responses.

This in turn may have palpable impacts on health outcomes, by providing quicker logistical relief to health services, or ensuring faster lockdown measures to reduce infection rates. This is highly salient information for anyone looking at policy reform to optimize civil-military operations. Militaries with policy-mandated public health operations within their military code may also have been more inclined to invest in training and education pertaining to public health, which in turn may have impacts on health outcomes during the pandemic, or impacted their success in maintaining order, instilling trust in government, enforcing lockdown measures, and aiding in recovery efforts. Policies pertaining to guardrails around civil-military operations may also inform research into authoritarianism escalation risk. Elucidating these questions will be crucial in preparing for future pandemic efforts.

The methodology employed in this research has important limitations. The protocol relied on open-source policies for included countries. As such, nations without freely accessible digital copies of its policies and legislation may not have been accurately represented. Furthermore, military operations may include a scope of practice not codified into policy. As such, some militaries may consider MOOTW within their remit, despite this not being reflected in military policy. In this way, our research represents a policy landscape, but may not represent the reality of domestic military operations. Finally, in periods of exceptional circumstances countries may deviate from their established policies through protocolised emergency powers, which have not been captured by this research.

## Conclusion

The data contained within this descriptive paper demonstrates the variety of ways in which countries define and regulate their military’s authority, operationalisation and the scope of domestic MOOTW. We applied this to the context of public health emergencies, which has become highly relevant since the mass deployment of militaries around the world in domestic public health responses. While we describe policies which were broadly drafted in non-emergency times, instances of deviation from these laws during public health emergencies were beyond the scope of our paper. In describing the present policies of civil-military operations, we generated and informed future research avenues on the military deployment for public health emergencies. This research therefore provides much needed data to generate evidence-based policy analysis for pandemic preparedness.

## Data Availability

All relevant data is available in a public open access repository. Data can be found on GitHub at https://github.com/cghss/military_engagement

## Acknowledgements

This research was funded by a grant from Rockefeller Foundation. The funder had no part in the conceptualization, design or analysis of the study. The authors would like to acknowledge the work of Dr Ellie Graeden, Tess Stevens, Hailey Robertson, and Ryan Zimmerman in contributing to designing of the data structure, managing of the project database, and quality control of the collected data.

